# Sleep duration and mental well-being are associated with delayed memory in a 12-week digital and remote community-sample study

**DOI:** 10.64898/2026.07.29.26359263

**Authors:** Sofia Marcolini, Sarah E. Polk, Emrah Düzel, David Berron

## Abstract

Unsupervised digital memory assessments are increasingly used in the field of Alzheimer’s disease to detect cognitive changes, recruit for clinical trials, and monitor cognitive changes longitudinally. However, the impact of factors like sleep and mental well-being on delayed memory task performance remains unclear. Adults aged 18 and older from a community sample across Germany (19-85 years) completed weekly a delayed memory recall task (Object-in-Room Recall, ORR) via the *neotivTrials* platform over 12 weeks. They also completed brief surveys on wake- and bedtime, and mental well-being (score created from happiness, calmness, energy, freshness, and interest items). Multilevel linear mixed models examined within- and between-person effects of sleep duration the night before testing (*n* = 329) and mental well-being over the preceding eight days (*n* = 234) on delayed associative memory. A quadratic term for sleep was included to test a hypothesized U-shape relationship, and interactions with age, gender, and subjective memory decline were analyzed. Sleeping more than one’s usual average the night before testing was associated with better delayed memory (*β =* 0.03, *p =* 0.02), while no between-person sleep effects emerged. Better delayed memory performance was also linked to better mental well-being, both relative to one’s own average (*β =* 0.05, *p =* 0.01) and the sample average (*β =* 0.12, *p =* 0.01). These effects were consistent across age, gender, and subjective memory decline, except for a stronger mental well-being-performance association at older ages. No quadratic effects of within- or between-person sleep duration on delayed memory were found. These findings suggest that participants’ averages of sleep duration and mental well-being should be considered when interpreting performance on remote digital memory assessments. Additionally, by enabling frequent assessments, remote testing designs allow the examination of within-person associations between predictors and cognition, unlike cross-sectional assessments, which rely on potentially noisy one-time estimates.

**Highlights:**

- Sleeping longer than one’s typical average relates to better delayed memory performance.
- Better mental well-being, relative to individual and group averages, is linked to better delayed memory.
- Sleep duration and mental well-being should be accounted for when interpreting results from digital memory testing.

**Author Summary:** Remote digital assessments of memory are increasingly being used to identify individuals at higher risk for Alzheimer’s disease, support clinical trial recruitment, and monitor cognitive changes over time. These tests are completed in naturalistic settings and are repeated frequently, providing a practical way to capture memory performance in everyday life. However, they are performed in unsupervised settings, and it is unclear how factors like sleep or mental well-being influence performance on such tests. To explore this, we asked adults from the community to take memory tests online for over 12 weeks and complete short surveys about their sleep duration and how they were feeling. We found that people had better memory after sleeping more than they usually did. Feeling mentally better, both compared with one’s own usual state and compared to others, was also linked to better memory. These patterns were similar across age, gender, and self-reported memory concerns, though mental well-being had a stronger effect for older adults. The findings highlight the importance of considering both sleep and psychological well-being when interpreting memory test results collected remotely. Moreover, they highlight how longitudinal designs allow the investigation of how individual’s factors influence memory over time by tracking day-to-day changes.

## Introduction

Digital cognitive testing is increasingly implemented to screen individuals at risk for Alzheimer’s disease (AD) and is starting to be considered in the recruitment and cognitive monitoring of clinical trials [1]. Remote and unsupervised testing has proven feasible and valid in older individuals with and without clinically established cognitive impairment [2,3] and offers advantages by enabling frequent and ecologically valid testing [4]. However, performance can be influenced by both external factors, such as environmental distractions, as well as internal factors, including sleep and mental well-being [5,6], which would be typically controlled for during in-clinic assessments by the assessor [5,7]. While the effects of some of these factors on several cognitive functions have been examined in recent studies [5,6,8], the influence of sleep duration and mental well-being on delayed memory assessed remotely in community-samples remains unexplored.

Delayed memory recall is particularly relevant in the context of cognitive decline as it is among the earliest cognitive functions disrupted in AD and constitutes a core criterion in its clinical diagnosis [9,10]. Performance on delayed memory recall tasks reflects the ability to consolidate and retrieve information over time, engaging hippocampal-dependent processes which are critical for memory formation [9–11]. Digital tools have become increasingly capable and advantageous in assessing these specific memory processes [12], also considering that recall and recognition tasks with longer delays of hours or days are difficult in in-clinic settings. Despite this, it remains essential to identify the internal and external factors that may influence performance to interpret results in context.

Sleep is a well-established contributor to cognitive function, with evidence suggesting a U-shaped relationship, meaning that both shorter and longer sleep relative to seven to eight hours is associated with poorer outcomes [13–15]. Studies examining associations between sleep and cognitive performance in naturalistic settings have predominantly focused on processing speed and working memory (see Supplementary Table 1 for an overview) [8,16–21]. Across community-dwelling middle-aged and older adult samples, findings suggest that within-person deviations from an individual’s typical sleep are associated with poorer next-day cognitive performance, whereas between-person differences in average sleep are often weak or absent [8,16,17]. Notably, despite this growing evidence, associations with delayed memory outcomes remain unknown.

Similarly, delayed memory assessed remotely remains unexplored in relation to mental well-being, namely the presence of positive psychological well-being and vitality, including being cheerful, calm, active, rested, and interested in daily life [22], which has been found to be consistently related to cognition in in-person designs [23–26]. The World Health Organization has developed a questionnaire to assess subjective mental well-being [27], and while it has been largely employed in various contexts and disease conditions, studies on its relationship with cognition are missing [28]. Using another mental well-being scale, higher levels of mental well-being were found to be associated with better in-person cognitive performance in a large community study [29]. Negative affect has been linked to slower digital survey completion [30], while better-than-average mood to improved working memory in adults under 65 years of age [6].

Since sleep and mental well-being fluctuate over time and often in person-specific ways [31–34], temporal and within-person effects cannot be disregarded. Remote digital testing facilitates frequent repeated assessments, allowing the examination of how temporal variations in internal factors relate to performance, which is essential to determine whether such factors should be accounted for when interpreting unsupervised test results.

In the present Citizen Science study, unsupervised digital memory tests were administered alongside brief weekly surveys over 12 weeks in a community sample of adults aged 18 years and older. The study aims to examine both within- and between-person effects of sleep duration and mental well-being on delayed memory performance, while adjusting for demographic variables and reported distractions during testing. We hypothesized that longer sleep duration or a U-shaped sleep duration pattern, along with better mental well-being, would predict better delayed associative memory performance, with these effects potentially moderated by age, gender, and subjective memory decline. Subjective memory decline was included as a moderator because prior work suggests that people with greater self-perceived cognitive concerns show a stronger link between poor sleep and objective memory decline, and that mood-related factors similarly shape how subjective and objective memory relate to one another [35,36].

Previous work suggests that processing speed and executive function may be more closely related to depressive symptoms in older than in younger adults [37] and that older adults may be more resistant to the acute effects of sleep deprivation than younger adults [38–40]. Therefore, we hypothesized that delayed associative memory would show stronger associations with sleep in younger adults, and with mental well-being in older adults. While results are mixed [41,42], we might expect stronger associations of sleep duration and mental well-being on cognition in women than men and stronger associations in those with subjective memory decline.

## Methods

### Study recruitment, procedures, and participants

The study was promoted through the online Citizen Science platform BürgerSchaffenWissen (www.buergerschaffenwissen.de), which receives support from the German Ministry of Research and Education, and additionally through the project website and local advertisements. Approval from the ethics committee at Otto von Guericke University Magdeburg was obtained. Participants were all 18 or older and provided informed consent. Detailed study procedures can be consulted elsewhere [43]. Briefly, interested individuals were directed to register for the study and download the *neotivTrials* app on their smartphones or tables (iOS or Android devices). Over the following 12 weeks, participants completed weekly memory assessments. Following each memory assessment, participants rated their own performance and concentration levels using a 5-point Likert scale (1 = very bad to 5 = very good) and indicated whether they had experienced any distractions during the task (yes/no). Alongside the memory assessments, participants filled out brief weekly surveys covering sleep duration and mental well-being.

### Measures

#### Demographics and memory task

Information on age and gender was obtained. Subjective memory decline was also tested asking participants’ judgment of whether their memory capacity had become worse during the 5 years preceding the study. The score was then categorized in stable or declining. In the current study, only one (Object-in-Room Recall [ORR] test) of the three memory tasks administered in the original study was used for analyses [43], due to its relevance for delayed associative memory and because of the largest sample availability. The ORR was developed to quantify hippocampal pattern completion, or the neural process by which partial memories are completed from earlier experienced episodes, which relies on the hippocampal *cornu ammonis* 3 (CA3); successful pattern completion supports associative memory [11]. In the ORR, participants learned the spatial positions of two objects in a room. During immediate recall (phase one), they were shown the empty room with a marked spot and had to choose the correct objects from three options: the target object, a distractor from the same room but wrong position, and a distractor from another room. After 24 hours, they completed an identical task (phase two), testing delayed recall with a randomized stimulus order. The 24-h delayed retrieval accuracy was calculated and used as outcomes of analyses.

#### Mental well-being

To measure mental well-being, the World Health Organization-Five Well-Being Index (WHO-5) [27] was administered, which is a five-item self-report instrument (items reported in Table 1). Questions referred to the eight days preceding the assessment, and answers were given on a scale from 0 = at no time to 5 = all of the time. The total score, ranging from 0–25, is calculated by summing the scores on each of the five questions. A raw score below 12 has been previously suggested as a cut-off for poor mental well-being and additional assessment for the possible presence of a mental health condition [22]. Results on this categorization are reported for descriptive purposes in Table 2. The WHO-5 questionnaire also shows a strong correlation with other established mental health measures like the Geriatric Depression Scale (4 and 15-item versions), Major Depression Inventory (MDI), Beck Depression Inventory Second Edition (BDI-II), and Patient Health Questionnaire-9 (PHQ-9) [44–46].

**Table 1.** Items from the World Health Organization-Five Well-Being Index (WHO-5) [27].

| Item | Description |
| --- | --- |
| **Happiness** | In the last 8 days I was happy and in a good mood. |
| **Calmness** | For the last 8 days I have felt relaxed and at ease. |
| **Energy** | In the last 8 days I have felt energetic and active. |
| **Freshness** | For the last 8 days I have felt fresh and rested when I woke up. |
| **Interest** | In the last 8 days my everyday life has been full of things that interest me. |

*Note.* Response levels: 0 = All the time; 1 = Mostly; 2 = Slightly more than half the time; 3 = Slightly less than half the time; 4 = From time to time; 5 = At no time point. The items reported in this table were paraphrased from the original items. Participants were presented the items translated in German.

**Table 2.** Sample characteristics.

|  | Sleep sample N = 329 | Mental well-being sample N = 234 |
| --- | --- | --- |
| Variable | Mean (Standard deviation), [Range] or N (%) |  |
| Age | 57.95 (12.12), [19.00 – 85.00] | 59.16 (11.62), [19.00 – 82.00] |
| Gender (female) | 254 (77.2 %) | 181 (77.4%) |
| Subjective memory decline |  |  |
| Stable | 196 (59.6 %) | 150 (64.1 %) |
| Declining | 68 (20.7 %) | 49 (20.9 %) |
| Missing data | 65 (19.8 %) | 35 (15 %) |
| Hours of sleep night before | 7.39 (0.88), [4.37 – 9.44] |  |
| Mental well-being (WHO-5 score) |  | 17.25 (3.99), [2.50 – 24.93] |
| Mental well-being* (poor) |  | 33 (14 %) |
| Subjective performance (1: very bad – 5: very good) | 2.61 (0.63), [1.00 – 4.18] | 2.64 (0.67), [1.00 – 4.25] |
| Concentration (1: very bad – 5: very good) | 3.67 (0.59), [1.50 – 5.00] | 3.70 (0.64), [1.67 – 5.00] |
| Distraction (yes) | 31 (9.4 %) | 23 (9.8 %) |
| Screen size | 14.14 (4.49), [10.16 – 32.85] | 14.25 (4.62), [10.16 – 32.85] |
| Score total (ORR) | 38.60 (3.34), [28.17 – 49.00] | 38.59 (3.60), [28.25 – 48.67] |

Note. ORR = Object-in-Room-Recall; \*most common answer per participant

#### Sleep

After completing phase one of the memory tasks (immediate recall), participants were asked to complete brief surveys which also included questions on sleep. In this study, we considered answers to the questions about the time they went to bed the night before and the time they woke up on the day of cognitive assessment. From these self-reported times, the number of hours of sleep the night before was calculated. Participants were also asked about the average hours of sleep in the past eight days. We only used the latter variable for descriptive purposes (Supplementary Fig S2), since we were primarily interested in the times participants went to bed and woke up the night before the test.

### Data cleaning and inclusion

Participants were included in the analyses if they provided valid information about their gender and age (≥18 years). To ensure sufficient data quality, we also excluded individuals that did not meet certain careless criteria. These are specified in detail in Fig 1 and in a pre-processing report (Supplementary file preproc-orr.html). Specifically, for the delayed memory task, participants were required to have used screens with a non-zero size. To ensure task engagement, individuals were included only if they had no more than 8 timed out trials in either phase one or two. Because there was a time lag between the memory assessments and the WHO-5 questionnaire, we only retained data points with a 5- to 11-day interval between the two. For both the predictors and the outcome we excluded outliers; specifically, only observations within ± 3.5 standard deviations from the mean were retained. Finally, we only included participants with at least three valid entries for both the predictors and delayed memory to ensure sufficient longitudinal data for the mixed-effects models.

**Figure 1.**
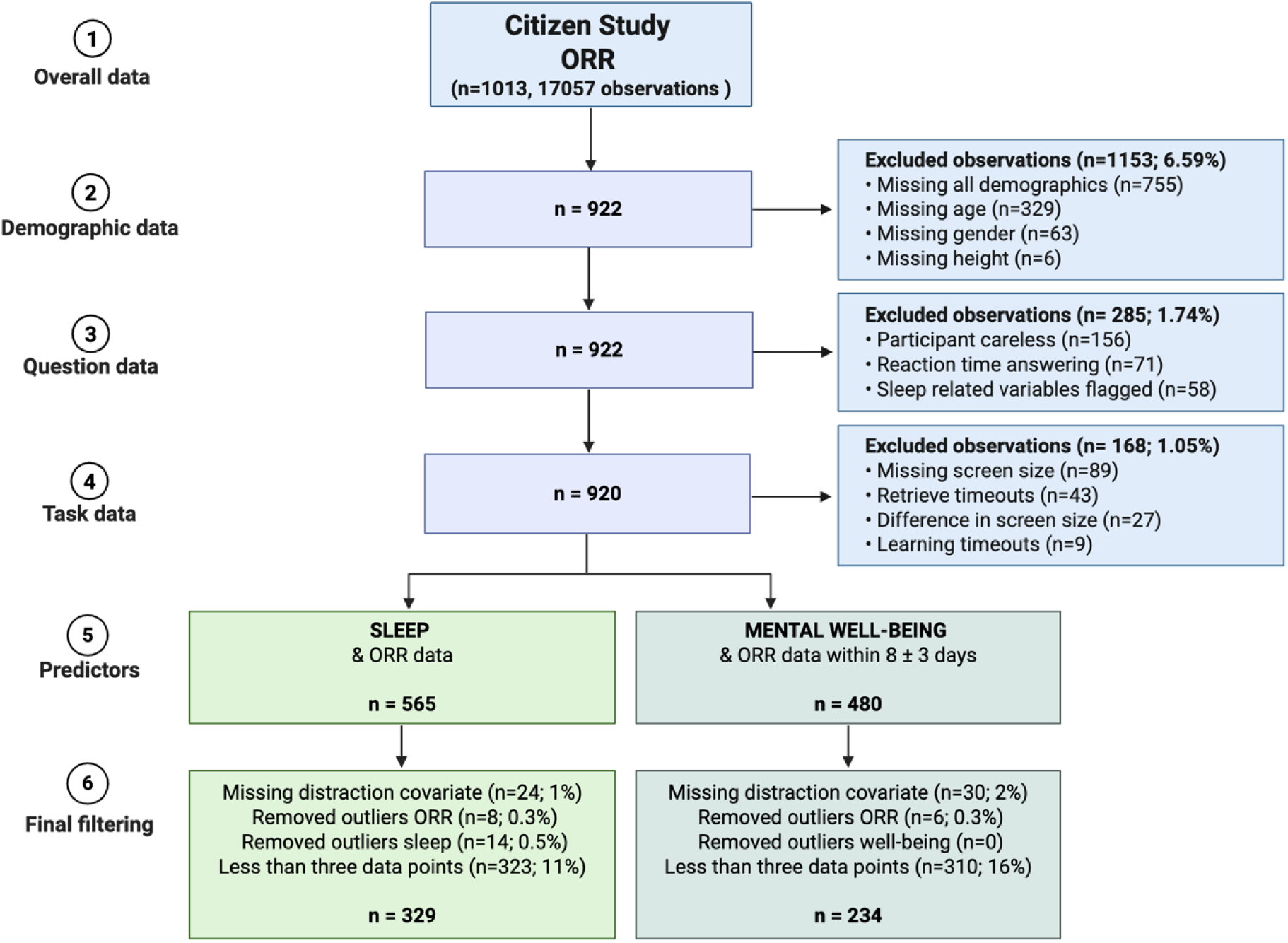
Inclusion flow. *Note.* ORR = Object-in-Room-Recall; Criteria for participant carelessness are described in the pre-processing report (Supplementary file preproc-orr.html).

### Data analysis

Data preparation and descriptive statistics were conducted in R 4.3.2 (R Core Team, 2023; see Supplementary Materials for all analyses code). An alpha of 0.05 was used to test statistical significance. To assess whether sleep and mental well-being were related to delayed memory over time, we ran multilevel mixed models using the ‘nlme’ package [47,48], which are well suited to the nested structure of longitudinal data [49]. The ICC for the score total of the ORR, which was calculated on 1594 observations, was 0.53 for the sleep dataset and 0.56 for the mental well-being dataset, meaning that 53% and 56% of the variance observed in the ORR was between people. Accordingly, two-level multilevel mixed models were computed, with observations (level 1) nested within participants (level 2). In the models, within- and between-person associations were separated [50–52]. Specifically, person-mean centering was used to index individual’s variation around their own mean (i.e., within-person variability). This is computed by first calculating each participant’s mean hours of sleep the night before and mental well-being across all observations. Next, the participant’s person-mean was subtracted from each observation to yield a person-mean-centered score for each measurement occasion. Conversely, grand-mean centering was used to index between-person variability in overall (time-invariant) sleep and mental well-being scores. This was computed by subtracting the sample’s grand mean score of sleep and mental well-being from each participant’s overall sleep and mental well-being mean score. This yielded a time-invariant grand-mean centered sleep and mental well-being for each participant. Time-varying person-mean centered (within-persons) and time-invariant person-mean (between-persons) sleep and mental well-being variables were then entered as independent variables in the model.

Two models were run for each dataset, specifically two models using the sleep within- and between-person predictors and two using the mental well-being within- and between-person predictors. Both Models 1 and 2 used the ORR as outcome. Models 1 were corrected for age and gender; Models 2 were additionally corrected for distractions. Average effects of sleep and mental well-being were estimated as fixed effects, with individual differences in baseline performance modeled as random intercepts. All models were fitted using restricted maximum likelihood and included random intercepts.

Using the covariates of Models 2, Models 3–5 tested interactions with age, gender, and subjective memory decline. Additionally, models inserting a quadratic term for the sleep predictors were also run to test for a U-shape relationship between sleep and memory. A sensitivity analysis was also conducted running analyses on the full sample without excluding outliers (which were for both the predictors and the outcome observations outside ± 3.5 standard deviations from the mean). Sleep variability indices have also been calculated for descriptive purposes, specifically the standard deviation and root mean square of successive differences (RMSSD; indicative of night-to-night changes).

## Results

The 329 participants included in the sleep analyses were on average 57.95 (± 12.12) years old and predominantly self-identified as female (77.2%); participants had between 3 to 16 assessments per person (mean = 7.88, SD = 3.86). The 234 participants included in the mental well-being analyses were 59.16 (± 11.62) years old on average and predominantly self-identified as female (77.4%); participants had between 3 to 15 assessments per person (6.65 ± 3.13). An overview of participant characteristics is provided in Table 2 and an inclusion flowchart in Fig 1. In Supplementary Fig 1, sleep variability indices are displayed showing that in this sample sleep duration is adequate (7.39 ± 0.88 hours per night) with sleep schedules being moderately irregular, with substantial inconsistency from night to night (RMSSD = 1.14). In Supplementary Fig 2, we provide a visualization of scatterplots and distributions between hours slept the night before, average hours slept per night the eight days before, and age.

### Within-person increases in self-reported sleep duration is associated with better delayed memory performance

Results from the multilevel models (sample including 1594 observations) are summarized in Table 3, including model fit comparisons. When accounting for age and gender, at the between-person level, hours of sleep the night before were not significantly related to delayed memory (*B =* 0.011, *p =* 0.762). At the within-person level, a positive association was found between hours of sleep the night before and delayed memory (*B =* 0.031, *p =* 0.023). This indicates that participants performed better on days following nights when they slept more than their own typical amount, whereas average differences in sleep across participants were unrelated to performance (Fig 2). Adding distraction as a covariate resulted in a modest but statistically significant improvement in model fit based on a likelihood ratio test (χ²(1) = 3.88, p = .049). No interactions were present with age, gender, nor subjective memory decline (Models 3 to 5; *p >* .05). In Fig 3, we show individual trajectories of memory performance and sleep in six exemplary participants. No quadratic effect was found between both within- and between-person sleep and memory (Supplementary Table 2).

**Figure 2.**
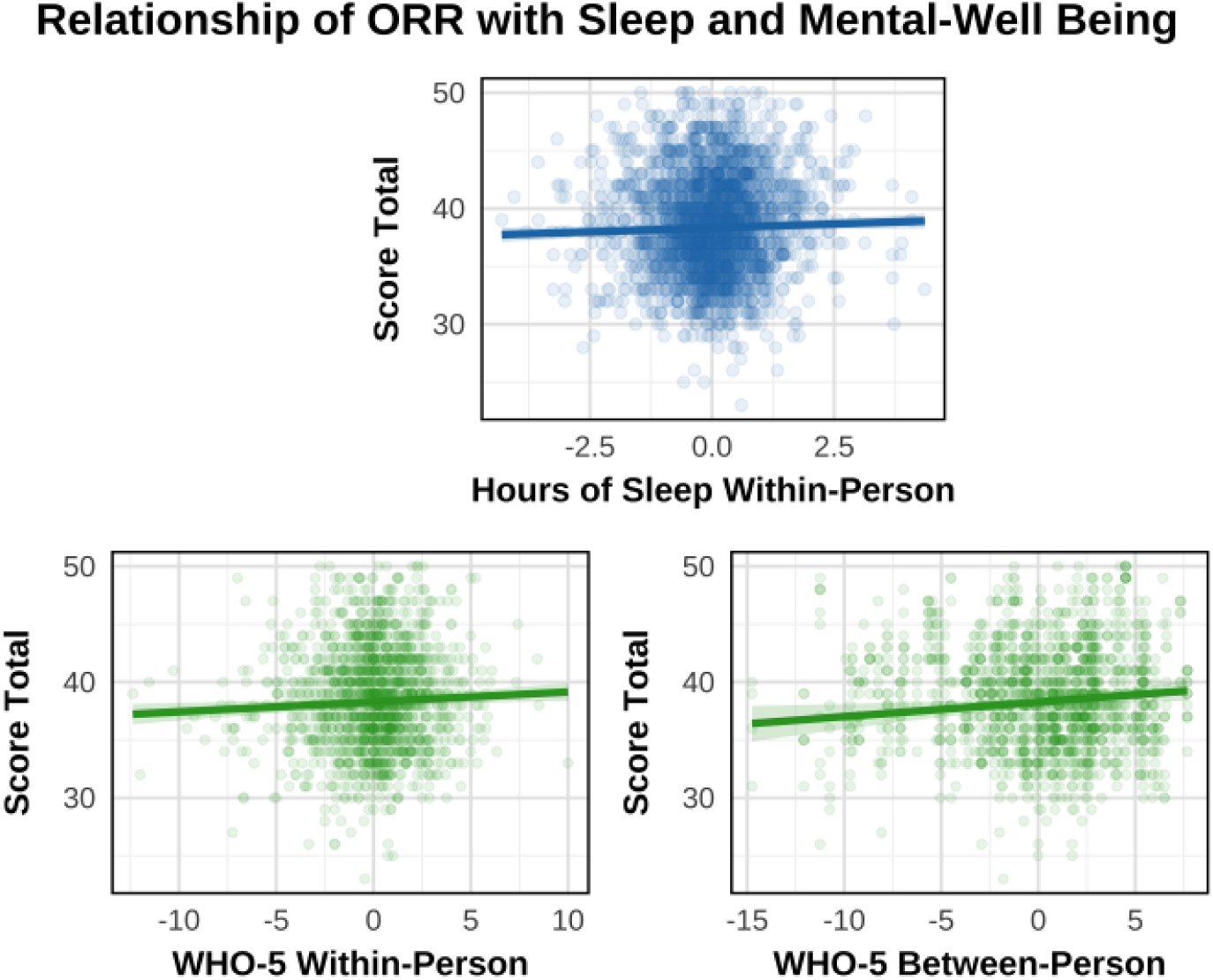
Associations between within-person mean hours of sleep the night before and within- and between-person mean mental well-being with delayed memory. *Note.* ORR = Object-in-Room-Recall; WHO = World Health Organization.

**Figure 3.**
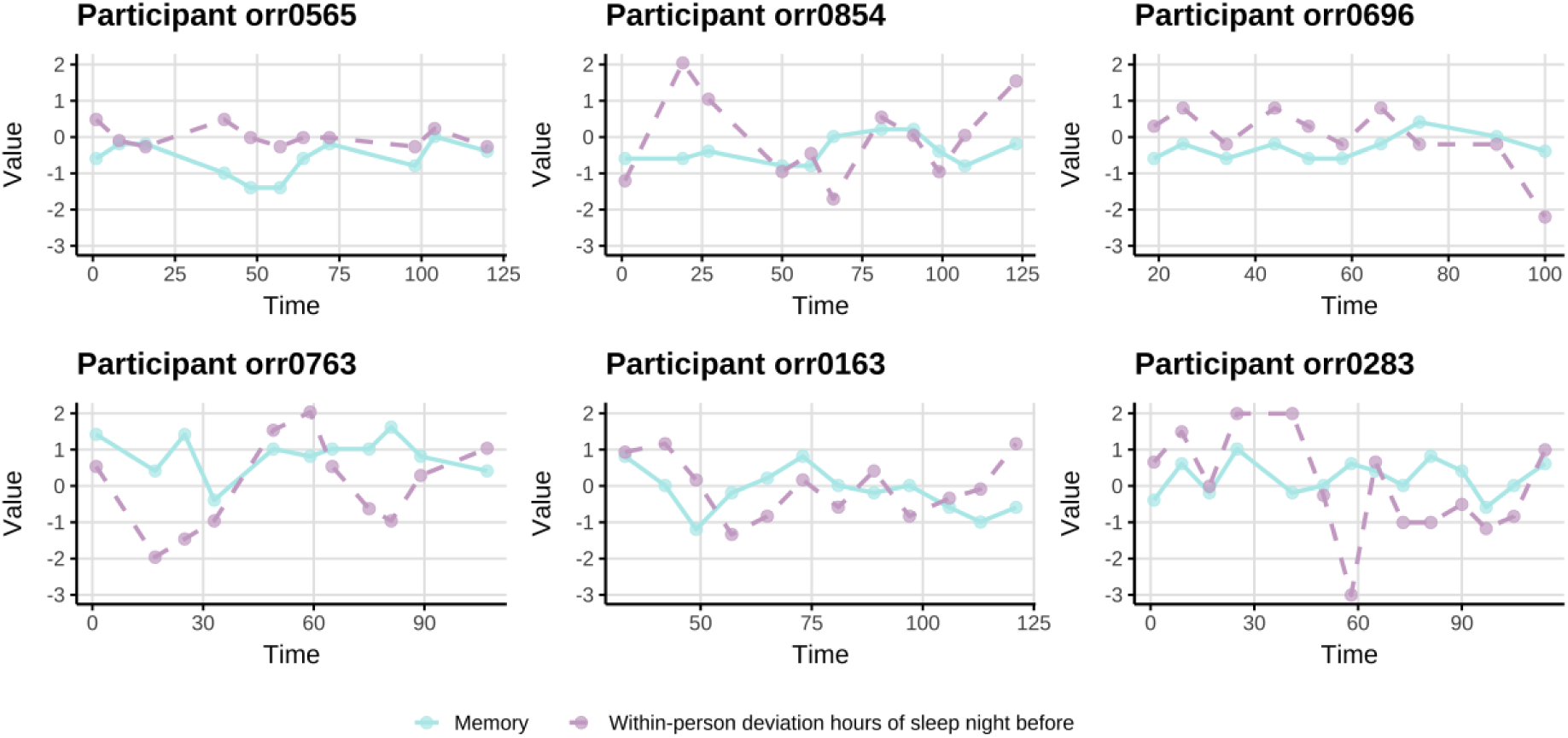
ORR performance and within-person deviation in hours of sleep the night before in six exemplary participants across time. *Note.* Individual trajectories are shown for six participants who’s within-person slopes of the z-scored value of score total (ORR outcome) on the within-person predictors of hours of sleep the night before, were closest to the group-level slope (0.024), among those with at least 10 observations. This selection highlights participants whose patterns are most representative of the overall trend.

**Table 3.** Within and between-person associations of hours of sleep and delayed memory and interaction models.

| | $\beta$ (std.) | SE | CI lower | CI upper | t-value | p |
| --- | --- | --- | --- | --- | --- | --- |
| **Model 1: Age + Gender** |  |  |  |  |  |  |
| Intercept | -0.011 | 0.043 | -0.095 | 0.073 | -0.256 | 0.798 |
| Hours of sleep within-person | 0.031 | 0.013 | 0.004 | 0.057 | 2.283 | **0.023** |
| Hours of sleep between-person | 0.011 | 0.036 | -0.060 | 0.082 | 0.303 | 0.762 |
| Age centered | -0.375 | 0.036 | -0.446 | -0.305 | -10.498 | **0.000** |
| Gender (male) | -0.125 | 0.089 | -0.300 | 0.050 | -1.401 | 0.162 |
| **Model 2: Age + Gender + Distraction** |  |  |  |  |  |  |
| Intercept | 0.005 | 0.043 | -0.080 | 0.090 | 0.117 | 0.907 |
| Hours of sleep within-person | 0.032 | 0.013 | 0.006 | 0.059 | 2.377 | **0.018** |
| Hours of sleep between-person | 0.008 | 0.036 | -0.063 | 0.080 | 0.230 | 0.818 |
| Age centered | -0.378 | 0.036 | -0.449 | -0.307 | -10.534 | **0.000** |
| Gender (male) | -0.125 | 0.089 | -0.301 | 0.051 | -1.402 | 0.162 |
| Distracted (yes) | -0.107 | 0.046 | -0.198 | -0.016 | -2.301 | **0.021** |
| **Model 3: M2 + interaction with age centered** |  |  |  |  |  |  |
| Intercept | 0.005 | 0.043 | -0.080 | 0.090 | 0.117 | 0.907 |
| Hours of sleep within-person | 0.031 | 0.014 | 0.004 | 0.058 | 2.256 | **0.024** |
| Hours of sleep between-person | 0.008 | 0.036 | -0.063 | 0.080 | 0.230 | 0.818 |
| Age centered | -0.378 | 0.036 | -0.449 | -0.307 | -10.534 | **0.000** |
| Gender (male) | -0.125 | 0.089 | -0.301 | 0.051 | -1.402 | 0.162 |
| Distracted (yes) | -0.107 | 0.046 | -0.198 | -0.016 | -2.298 | **0.022** |
| Hours of sleep within-person* Age centered | -0.004 | 0.012 | -0.028 | 0.021 | -0.285 | 0.775 |
| **Model 4: M2 + interaction with gender** |  |  |  |  |  |  |
| Intercept | 0.005 | 0.043 | -0.080 | 0.090 | 0.115 | 0.909 |
| Hours of sleep within-person | 0.042 | 0.016 | 0.012 | 0.073 | 2.717 | **0.007** |
| Hours of sleep between-person | 0.008 | 0.036 | -0.063 | 0.080 | 0.230 | 0.818 |
| Gender (male) | -0.125 | 0.089 | -0.301 | 0.051 | -1.402 | 0.162 |
| Age centered | -0.378 | 0.036 | -0.449 | -0.307 | -10.534 | **0.000** |
| Distracted (yes) | -0.106 | 0.046 | -0.197 | -0.015 | -2.287 | **0.022** |
| Hours of sleep within-person* Gender (male) | -0.041 | 0.031 | -0.102 | 0.020 | -1.318 | 0.188 |
| **Model 5: M2 + interaction with subjective memory** |  |  |  |  |  |  |
| Intercept | 0.028 | 0.054 | -0.077 | 0.133 | 0.521 | 0.602 |
| Hours of sleep within-person | 0.037 | 0.017 | 0.004 | 0.070 | 2.200 | **0.028** |
| Hours of sleep between-person | 0.031 | 0.040 | -0.048 | 0.110 | 0.775 | 0.439 |
| Subjective memory decline (yes) | -0.081 | 0.094 | -0.266 | 0.104 | -0.862 | 0.389 |
| Age centered | -0.391 | 0.040 | -0.470 | -0.312 | -9.769 | **0.000** |
| Gender (male) | -0.111 | 0.098 | -0.304 | 0.083 | -1.127 | 0.261 |
| Distracted (yes) | -0.098 | 0.049 | -0.195 | -0.001 | -1.976 | **0.048** |
| Hours of sleep within-person* Subjective memory decline (yes) | -0.021 | 0.035 | -0.091 | 0.048 | -0.596 | 0.551 |

Note. Standardized coefficients ( $\beta$ ) reported. CI = 95% confidence interval; bold p-values are < .05. Model fit statistics: Model 1 (Age + Gender): AIC = 13596.07, BIC
= 13637.08, logLik = -6791.03; Model 2 (Age + Gender + Distractions): AIC = 13594.18, BIC = 13641.05, logLik = -6789.09; Model 3 (Interaction Age): AIC = 13604.93,
BIC = 13657.66, logLik = -6793.47; Model 4 (Interaction Gender): AIC = 13596.56, BIC = 13649.28, logLik = -6789.28; Model 5 (Interaction Subjective memory decline):
AIC = 11123.9, BIC = 11180.51, logLik = -5551.95.

### Within- and between-person self-reported increase in mental well-being is associated with better delayed associative memory performance

Results from the multilevel models are summarized in Table 4 (sample including 1557 observations), including model fit comparisons. When accounting for age and gender, a positive association was found between mental well-being of the past eight days and delayed memory at both the within-person (*B =* 0.047, *p =* 0.007) and at the between-person (*B =* 0.116, *p =* 0.012) level. This indicates that participants with higher overall levels of well-being, as well as those experiencing better-than-usual well-being, performed better on delayed memory tasks (Fig 2). Model comparison suggested that including distraction as a covariate did not substantially improve model fit (likelihood ratio test: χ²(1) = 0.22, p = .64). No interactions were found with gender, nor subjective memory decline (Models 4 and 5; *p >* .05), although a significant interaction was present with age, showing stronger effects of mental well-being within-person on performance with increasing age (Table 4 Model 3; Supplementary Fig 3). Sensitivity analyses showed that results remain unvaried when using the sample without excluding outliers (Supplementary Table 3 and 4).

**Table 4.**
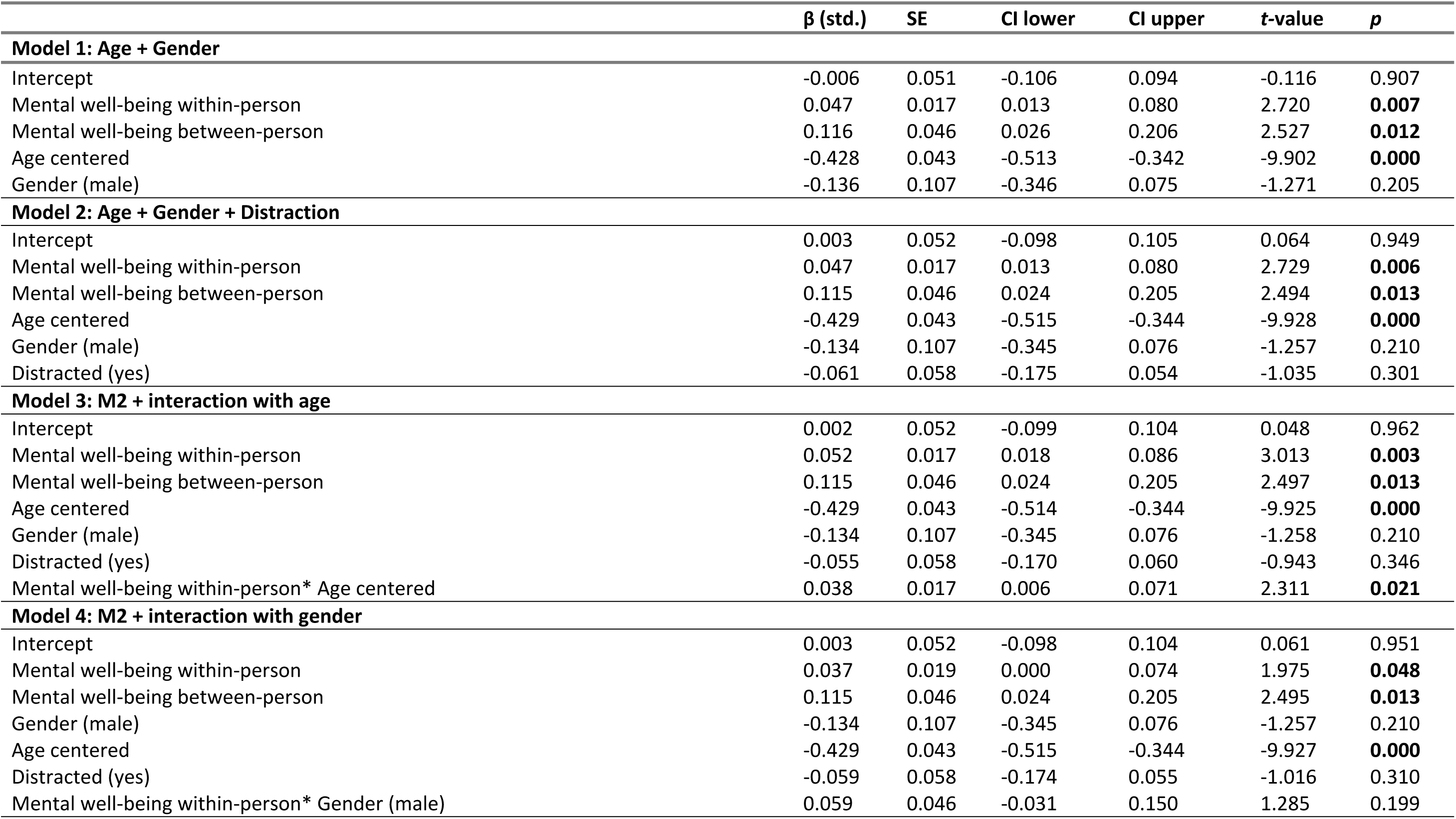
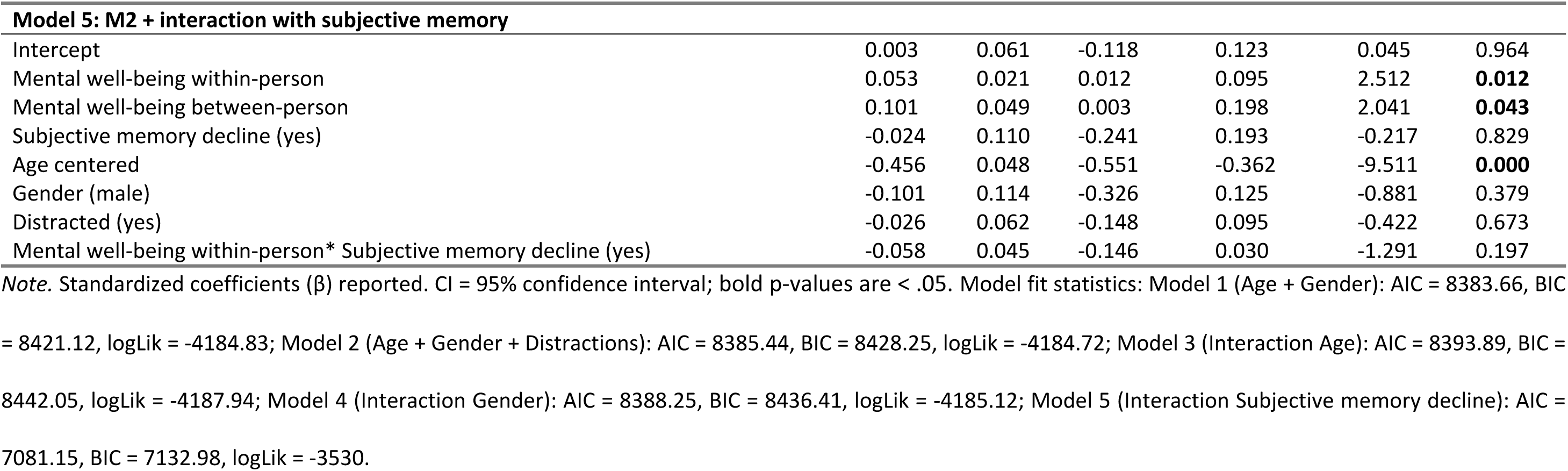
Within and between-person associations of mental well-being and delayed memory and interaction models.

## Discussion

In this 12-week Citizen Science study, we found that participants performed better on a digital and unsupervised delayed associative memory test when they slept more than their own average the night before testing, but not when they slept more than the sample average. Additionally, participants with higher overall levels of mental well-being performed better, as did those experiencing better-than-usual well-being, particularly among older participants. These results suggest that sleep and mental well-being should be taken into consideration when interpreting delayed associative memory tests performed remotely.

### Sleep and delayed associative memory

Meta-analytic evidence indicates that sleep restriction (3–6.5 hours) compared to normal sleep (7–11 hours) negatively affects memory formation, though with a small effect size [42]. In a large online study (*N* = 10,000), self-reported sleep duration over the past month showed a U-shaped association with verbal ability, reasoning, and overall cognitive performance, but not with short-term memory [13]. In our study, we do not find a U-shaped relationship between sleep duration and delayed memory assessed remotely, but rather a linear association indicating that sleeping more than one’s average is beneficial for delayed memory performance. Our results align with emerging evidence that within-person fluctuations in sleep are informative for understanding daily processing speed and working memory performance [8,16,17]. We extend these findings to delayed associative memory, a cognitive domain with a central role in memory consolidation and early vulnerability in Alzheimeŕs Disease (AD), suggesting that sleep-related cognitive benefits observed in digital paradigms are not limited to attentional or executive processes but also generalize to hippocampal-dependent memory consolidation.

In contrast with our results, Buxton et al. did not find an association between sleep duration and daily cognitive tests, but only with indices of sleep fragmentation (wake after sleep onset; WASO). The average age of their sample was 22 years older than ours and consisted of community-residing adults. Given that older individuals experience more WASO episodes, it is possible that sleep fragmentation, which we could not assess due to the absence of objective sleep measures, is more likely to influence cognition in older adults than sleep duration per se [54]. This could also explain the lack of age-moderating effects found in our study as well as in previous studies, showing that the relationship between sleep duration and cognition appeared invariant across adult age groups [17,18]. Together, these findings suggest that short-term sleep-related fluctuations in cognitive performance may operate similarly across adulthood, even as absolute levels of cognitive functioning decline with age.

Importantly, another factor that may have contributed to differences across studies is the consideration of environmental distractions during cognitive testing. Among previous studies (Supplementary Table 1), only one adjusted for distractions during task completion [17]. In our study, we accounted for self-reported distractions which were independently associated with poorer delayed associative memory performance. However, as the within-person association between sleep duration and delayed memory remained significant after adjusting for distraction, the observed sleep–memory relationship is unlikely to be explained solely by situational testing conditions.

Consistent with prior evidence, we did not observe between-person effects of sleep duration. This finding may reflect individual differences in sleep need and support the notion that the cognitive benefits of sleep are person specific [55]. In line with this interpretation, Wild et al. demonstrated that sleep “delta” (the difference between the sleep duration the night before testing and one’s usual sleep) was associated with overall cognitive performance, highlighting the importance of intra-individual variability rather than absolute sleep duration [13].

Our findings are also consistent with more traditional study designs. For example, an in-person cross-sectional study showed that sleeping more than usual, but not less than usual, the night before testing was positively associated with information processing speed [56]. However, cross-sectional assessments of “usual” sleep are susceptible to reporting bias, which can be mitigated by calculating within-person averages based on sleep duration reported across multiple days, as done in the present study. Similarly, a study examining the effects of prior-night sleep duration on cognition across eight testing sessions reported associations with sleep duration, although results were based on a global cognitive composite rather than specific cognitive domains and did not employ digital cognitive assessments [57].

### Mental well-being and delayed associative memory

Affect, or momentary emotional state, has been shown to influence cognitive performance [30], although evidence of this relationship in digital memory testing remains limited. Our findings indicate that higher mental well-being is associated with better delayed associative memory performance, and even stronger effects are observed for higher mental well-being compared to one’s own average.

Almost no studies have investigated the association between mental well-being and delayed associative memory. One population-based cohort study reported positive associations between psychological well-being and global cognitive function across multiple domains, including delayed memory, although cognition was not assessed digitally [29]. Findings from cross-sectional community samples are otherwise mixed, likely reflecting heterogeneity in how the concepts of mental well-being, affect, and mood are operationalized. Mood and affect refer to internal emotional states characterized by valence and arousal and reflect transient subjective experiences, whereas mental well-being represents a broader evaluation of psychological functioning and life circumstances.

Evidence regarding mood and affect specifically shows inconsistent associations with cognition. Lower mood has been linked to increased inhibitory control in a small sample (*N* = 106; [20]), while higher positive affect has been associated with greater effortful attention (*N* = 122; [21]). In contrast, no significant associations between mood and cognition were found in a larger sample (*N* = 2,018; [22]). More recently, negative affect has been shown to predict slower survey completion times, a proxy for cognitive processing speed (*N* = 914; [27]), and better-than-average mood was associated with improved working memory in individuals younger than 65 years [6]. However, most of this work focuses on between-person differences and does not capture within-person variability. Although, a recent Ecological Momentary Assessment study involving university students has shown that within person fluctuations in negative affect significantly predicted increased reaction time inconsistency, highlighting the role of emotional processes in cognitive performance [58].

Our study examines within-person fluctuations and assesses mental well-being over the eight days preceding testing. Given this time frame, focusing on mental well-being, a broader construct than momentary mood or affect, may be better suited to the study design. Nonetheless, additional research using digital memory assessments is needed to replicate and extend our findings on the relationship between mental well-being and delayed associative memory.

### Implications of findings and physiological underpinnings

These findings have practical implications for digital cognitive assessments. Collecting information on participants’ sleep the night before testing and their mental well-being can help contextualize performance and improve the interpretability of remote assessments, particularly in longitudinal and repeated-measures designs. In line with emerging evidence that arousal and environmental distractions during task completion can influence remotely assessed cognition [5], our results support that self-reported distractions should be considered when interpreting test scores.

Although not the primary focus of the present study, our findings may also inform the broader literature linking sleep and mental well-being to neurodegeneration and age-related cognitive decline. Sleep disruption has been extensively reported as a risk factor for AD [59,60]. Proposed mechanisms include increased neuronal activity during prolonged wakefulness, leading to elevated production and aggregation of amyloid-β, a central pathological hallmark of AD [61]. In addition, heightened sympathetic activity during wakefulness suppresses glymphatic system function, potentially impairing the clearance of neurotoxic proteins. Sleep loss is also associated with oxidative stress, inflammation, and disruptions in synaptic homeostasis, all of which may contribute to neurodegenerative processes [61]. Consistent with these mechanisms, short self-reported sleep duration has been associated with higher amyloid-β burden in older adults [62].

Mental well-being, while a broader and less well-defined construct in relation to neurodegeneration, has also been linked to cognitive aging, although much of the existing evidence is correlational [63]. Greater resilience and lower perceived stress appear to buffer the neurotoxic effects of chronic hypothalamic-pituitary-adrenal (HPA) axis activation and glucocorticoid exposure, which are implicated in hippocampal atrophy and other neurodegenerative changes [64]. In addition, better psychological well-being is associated with lower systemic inflammation and healthier lifestyle behaviors, both of which may reduce neuroinflammatory processes contributing to neuronal damage in AD and related dementias [65].

Integrating evidence from intensive longitudinal designs that capture within-person associations between internal states and cognition with longitudinal biomarker assessments may provide a powerful framework for examining how these relationships evolve across aging. Such an approach could inform intervention studies targeting sleep and mental well-being, helping to clarify their potential role in preventing or slowing neurodegeneration-related cognitive decline and in identifying the psychological mechanisms underlying intervention effects.

### Limitations and future directions

Several limitations should be considered. First, we did not collect participants’ educational attainment, precluding adjustment for this potentially important covariate. Although, in previous studies from our group, no effect of education on performance in this task was found [66,67]. Second, mental well-being was assessed retrospectively over the past eight days rather than momentarily, which may introduce recall bias. Although we minimized the interval between cognitive and well-being assessments, future studies should explore momentary assessments of mental well-being immediately prior to cognitive testing. Experience sampling designs with multiple daily assessments over consecutive days would enable cross-lagged analyses to examine bidirectional associations between sleep, mental well-being, and memory performance, potentially allowing for Granger-causal inference. In the present study, sleep and mental well-being were examined independently, despite being intrinsically related. Future work could apply dynamic structural equation modeling (DSEM) to disentangle their temporal interplay and to assess whether both constructs are required or whether one may serve as a proxy for general psychological well-being. Notably, subjective sleep reports, while not always aligned with objective sleep measures, are strongly influenced by emotional state, supporting their potential utility as proxies for mental well-being [68,69].

Third, we did not assess sleep quality or collect objective sleep measures (e.g., actigraphy), which could provide complementary information beyond sleep duration. Nevertheless, for the aims of the present study, asking brief questions on sleep before performance on cognitive tests is both feasible and minimally burdensome, which is an important consideration as cognitive assessments increasingly adopt high-frequency, short-duration digital designs. Fourth, future research should account for additional contextual factors during testing, such as social environment or physical location, which may influence cognitive performance and moderate associations between internal states and cognition.

Finally, as sleep duration represents a modifiable lifestyle factor [14], future research could consider collecting data on additional behaviors, such as physical activity or social engagement, to examine whether and how sleep influences cognition indirectly through individuals’ behavior. This approach may help elucidate mechanisms underlying sleep–cognition associations, as has been demonstrated in cross-sectional research linking physical activity to cognitive performance [70].

## Conclusion

In a Citizen Science study of 329 and 234 community-dwelling adults, we observed within-person associations between sleep duration and delayed associative memory, as well as within- and between-person associations between mental well-being and delayed associative memory. Our findings highlight the importance of considering sleep duration and mental well-being when interpreting digital memory assessments, and demonstrate that remote, frequent testing designs enable the examination of within-person associations between predictors and memory performance, rather than relying on single, potentially noisy estimates.

## Acknowledgements

We would like to thank all participants for taking part in the study, as well as to the Ministry of Labor, Social Affairs, Health and Equality (Minister Petra Grimm-Benne) for their patronage, and to the Minister-President of Saxony-Anhalt (Dr. Reiner Haseloff) for supporting the project. Our thanks also go to the Citizen Science Platform BürgerSchaffenWissen for providing us with the opportunity to promote our study to the wider public.

## Data availability statement

The minimal data set will be available on a repository with the associated DOI before publication. The code for data cleaning has been submitted as Supplementary Material.

